# Validity and Test-Retest Reliability of the Hume Pod Bioimpedance Analyzer for Body Composition Assessment

**DOI:** 10.64898/2026.07.17.26358337

**Authors:** Grant M. Tinsley, Carina M. Velasquez, Christine M. Florez, Ainsley E. Way, Madison H. Sullivan, Julia A. Whitson, Reva Ann Rudolph, John R. Alexander, Adhirath Malladi

## Abstract

Consumer-grade bioelectrical impedance analyzers have become widely used for body composition assessment, yet their accuracy varies considerably across devices. The Hume Pod is a popular consumer-grade analyzer marketed as being highly accurate, but independent validation is lacking. The purpose of this study was to evaluate the reliability and validity of the Hume Pod relative to both a four-compartment (4C) model and dual-energy X-ray absorptiometry (DXA). Sixty-seven adults (42 females, 25 males; age 37.2 ± 13.5 years, body mass index: 24.6 ± 4.9 kg/m^2^, body fat percentage [BF%]: 26.4 ± 10.2%) completed duplicate Hume Pod assessments alongside DXA and 4C evaluations. Reliability was evaluated using the technical error of measurement (TEM) and intraclass correlation coefficients (ICC). Validity was assessed using equivalence testing, Lin’s concordance correlation coefficient (CCC), standard error of the estimate (SEE), Bland-Altman analysis, and additional tests. The Hume Pod demonstrated strong reliability, with ICCs ≥ 0.993 and TEMs of 0.8% for BF% and 0.6 kg for fat mass (FM) and fat-free mass (FFM). Relative to the 4C model, BF%, FM, and FFM estimates were statistically equivalent (all *p*<0.05), with strong agreement (CCC=0.95-0.98), low SEE values (3.1%, 2.3 kg, and 2.2 kg, respectively), moderate limits of agreement (±6.1%, ±4.5 kg, and ±4.5 kg), and no proportional bias. Compared with DXA, generally strong agreement was also observed. These findings indicate that the Hume Pod demonstrates strong reliability and validity compared with laboratory reference methods for body composition estimation, supporting its potential use as a consumer body composition assessment.

## Introduction

Body composition assessment plays an important role in the evaluation of nutritional status, obesity management, clinical care, and healthy aging. Compared with anthropometric measures such as body mass index, assessment of fat mass (FM) and fat-free mass (FFM) provides a more physiologically meaningful characterization of nutritional status^(1)^ and allows clinicians, researchers, and individuals to better evaluate changes resulting from lifestyle or medical interventions.^(2; 3)^ As interest in body composition has expanded, there has been growing demand for assessment methods that are both accurate and practical for routine use. Although criterion methods such as multi-compartment models and magnetic resonance imaging provide highly accurate assessments, their cost, technical requirements, and limited accessibility restrict their routine application.^(4)^ Likewise, while dual-energy X-ray absorptiometry (DXA) is widely used in research and clinical practice,^(5)^ repeated laboratory assessments remain impractical for many individuals.

Bioelectrical impedance analysis (BIA) has become one of the most widely used approaches for body composition assessment because it is inexpensive, rapid, non-invasive, and well suited for repeated measurements outside of laboratory settings.^(6; 7)^ Continued advances in electronics, mobile applications, and analytical algorithms have accelerated the adoption of consumer-grade BIA devices, enabling individuals to monitor body composition at home with minimal burden. These developments have coincided with increased commercial interest in body composition analyzers. The global market for body composition analyzers has experienced substantial growth and is projected to expand from approximately USD 714 million in 2024 to USD 1.26 billion by 2033, with a substantial portion of this market being home use.^(8)^ Recently, the consumer bioimpedance market has expanded beyond conventional bathroom scales with basic body composition integration to include “premium” body composition analyzers that are marketed as providing laboratory-quality assessments through proprietary analytical methods. As these devices increasingly influence health, fitness, and weight-management decisions, their performance should be established through rigorous independent validation rather than manufacturer claims alone.

Despite their widespread use, the validity of consumer-grade BIA devices remains highly variable. While BIA analyzers generally rely on the same underlying physiological principles, differences in electrode configuration, measurement frequencies, signal processing, and proprietary prediction equations can produce substantially different estimates.^(7; 9; 10)^ Consequently, the performance of one BIA device cannot be assumed to represent that of another. In a previous investigation from our laboratory,^(11)^ the validity of fourteen consumer-grade BIA devices ranged from very poor to very good when evaluated against a criterion four-compartment (4C) model, demonstrating that considerable performance heterogeneity is present even among devices that appear similar and are intended for similar consumer applications. These findings underscore the importance of evaluating individual devices independently, particularly those that have achieved widespread adoption or claim superior analytical performance.

One such device that demonstrates substantial consumer popularity is the Hume Pod (Hume Health LLC, Wilmington, DE, USA). The Hume Pod is a portable, consumer-grade body composition analyzer that has gained notable popularity among fitness professionals and health-conscious consumers. Unlike many traditional single-frequency consumer BIA devices, the Hume Pod is marketed as an advanced system utilizing multi-frequency bioelectrical impedance and proprietary analytical algorithms to provide more accurate estimates of body composition and to serve as an alternative to laboratory-based assessment.^(12)^ Manufacturer-provided information also indicates that the Hume Pod body composition algorithms were independently evaluated against DXA data.^(13)^ Given its increasing use and positioning within the premium consumer body composition market,^(8)^ independent evaluation of the performance of this consumer-grade BIA analyzer is warranted. However, to our knowledge, no independent peer-reviewed validation of the Hume Pod relative to criterion body composition methods has previously been conducted.

Therefore, the purpose of the present study was to evaluate the validity and test-retest reliability of the Hume Pod for estimating body composition in adults relative to both a 4C model and DXA. The 4C model is widely regarded as the criterion field method for body composition assessment at the molecular level,^(14; 15)^ whereas DXA remains one of the most commonly used laboratory methods in research and clinical practice.^(5)^ Comparison with both reference methods provides a comprehensive evaluation of the performance of this increasingly popular consumer body composition analyzer. Based on our previous investigation of consumer-grade BIA devices,^(11)^ we hypothesized that the Hume Pod would demonstrate excellent test-retest reliability and strong linear associations with the reference methods but would produce estimates that significantly differ from reference methods.

## Experimental Methods

### Participants

Adults aged 18 to 75 years were recruited for participation from the local community in Lubbock, Texas, United States. Participants were required to be weight stable, defined as no change in body mass greater than 2.2 kg in the previous month, based on self-report. Individuals were ineligible for participation if they had a height >191.8 cm or weight >150 kg (due to device limitations), were unwilling to trim facial hair to ≤1.3 cm in length (due to the influence of hair on the air displacement plethysmography test), had a pacemaker or electrical implant (due to bioimpedance tests), had a prior amputation or physical deformity or abnormality that would invalidate body composition assessments, had a notable amount of implanted metal (e.g., full joint replacement), were claustrophobic, were unable to complete the overnight fast, or were currently pregnant or breastfeeding based on self-report. The study procedures were explained to all interested individuals, and each participant provided written informed consent prior to participation. This study was approved by the Texas Tech University Institutional Review Board (protocol #2025-614).

### Study Procedures

Participants reported to the research laboratory after an overnight (≥8-hour) period of abstention from food, drink, caffeine, nicotine, and dietary supplements. The evening prior to the research visit, each participant was asked to drink 1 liter of water after their final meal but before the beginning of the food and fluid abstention period to promote appropriate tissue hydration. Participants were additionally asked to abstain from exercise or other vigorous physical activities for ≥24 hours prior to their research visit. Upon confirmation that these pre-assessment restrictions were followed, participants changed into minimal form-fitting clothing for subsequent assessments.

Each participant voided their bladder and collected a mid-stream urine sample for assessment of hydration status, as urine specific gravity, via digital refractometer (PA201X-093, Misco). Height was measured to the nearest 0.1 cm using a mechanical stadiometer (HM200P, Charder Medical), and body mass (BM) was measured using a calibrated scale (Model BWB-627-A, modified Tanita Corp). Participants completed air displacement plethysmography (Bod Pod, Cosmed USA), dual-energy X-ray absorptiometry (DXA; iDXA, General Electric; enCore software v. 18 SP5), and professional-grade multi-frequency bioelectrical impedance analysis (mBCA 554 Ultra, Seca). The total body water (TBW) estimation equation used by this Seca bioimpedance analyzer was developed and validated using deuterium dilution^(16)^ and has previously demonstrated the highest accuracy of several bioimpedance analyzers for TBW estimation, as compared to deuterium dilution, in our laboratory (*data under review*). Data from the aforementioned assessments were used in the construction of a reference 4C model, with body volume (BV) taken from air displacement plethysmography, bone mineral content from DXA, TBW from the professional bioimpedance analyzer, and BM from the calibrated scale. After conversion of bone mineral content to bone mineral (Mo; Mo = BMC × 1.0436),^(17)^ these inputs were used for FM estimation within the 4C equation of Wang et al.^(14)^:

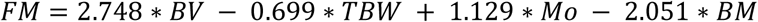

FM was subtracted from BM to yield an estimate of FFM, and FM was divided by BM and multiplied by 100 to generate body fat percentage (BF%). DXA-provided values of FM, FFM (i.e., lean soft tissue plus bone mineral content), and “region” total BF% were also used for the version of the analysis in which DXA served as the reference method.

During the same research visit, participants were assessed with the Hume Pod (Hume Health LLC), which was purchased online directly from the manufacturer’s website in January 2026. A new profile was created for each participant to ensure measurements remained independent of other participants. Within the accompanying software application, each participant’s sex, year of birth, and height were entered. Participants were then assessed two separate times separated by ∼10 minutes. For each assessment, the participant stepped on the scale, making contact between their feet and foot contact electrodes, and held the handle with hand contact electrodes slightly above waist level. After each successful test, results were saved for analysis. The “body fat mass” value was used for FM, and FFM was calculated as “weight” from the Hume Pod minus FM. BF% was calculated as FM divided by “weight” from the Hume Pod, multiplied by 100.

### Statistical Analysis

Due to the lack of prior peer-reviewed research on the Hume Pod, the sample size was informed by practical considerations and efforts to approximate the sample size of the prior consumer-grade bioimpedance work from our laboratory (n=73)^(11)^. The test-retest reliability of the Hume Pod was established from duplicate assessments performed within the single laboratory visit and therefore primarily reflected the technical error of the analyzer itself. The validity was separately examined using two criterion methods: the 4C model and DXA alone. The 4C model was implemented based on its known strengths for reducing the assumptions of FFM characteristics, while DXA was additionally employed due to its wide acceptance as a validated laboratory method, its greater availability as compared to a 4C model, and the prior use in the manufacturer’s reported evaluation of their analyzer.^(13)^

The reliability of the Hume Pod for BF%, FM, and FFM was evaluated through the absolute and relative technical error of the measurement (TEM). These were calculated as follows, where *V1* and *V2* are the first and second values obtained from repeated measurements for a particular variable, *n* is the sample size, and the grand mean is the mean of *V1* and *V2* means:

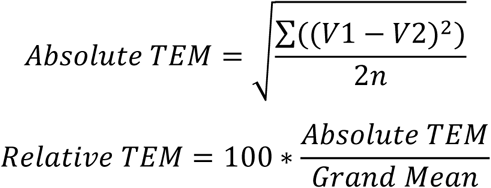

The absolute and relative TEM are synonymous with the precision error and root-mean-square coefficient of variation, respectively. The absolute TEM is presented in the unit of measurement for each variable, while the relative TEM is presented as a percentage. For BF%, only the absolute TEM was considered since this outcome is already presented in percentage units. The ICC was calculated using model 2,1.^(18; 19)^

The validity of Hume Pod body composition estimates was evaluated through separate comparisons with a 4C model and DXA. The linear relationships between the Hume Pod and each criterion method were established using ordinary least squares regression and comparison to the line of identity (*y* = 0 + 1*x*), with the reference method specified as the *y* variable and the Hume Pod specified as the *x* variable. To determine if values demonstrated group-level statistical equivalence with reference values, equivalence testing^(20)^ was performed using equivalence regions of ±2.0% for BF% and ±5.0% for FM and FFM.^(21)^ Paired-samples t-tests were also performed. The constant error (mean difference between methods) was calculated, along with the standard error of the estimate (SEE), root mean square error (RMSE), mean absolute error (MAE), Pearson’s *r* and R^2^, and Lin’s concordance correlation coefficient (CCC). Bland-Altman analysis was performed to establish the 95% limits of agreement, alongside linear regression to evaluate proportional bias.^(22)^ Statistical significance was accepted at *p*<0.05. All statistical analyses were conducted in R (v. 4.6.0).^(23)^

## Results

### Participants

Sixty-seven participants (42 F, 25 M) were included in the validity analysis (Table 1). Two individuals were excluded from the reliability analysis due to having only one valid test, resulting in a sample size of n=65 (41 F, 24 M) for the reliability analysis.

**Table 1.** Participant Characteristics. *Body composition values taken from the 4-compartment model; *F: female; M: male; M: mean; SD: standard deviation; USG: urine specific gravity*

|  | **All**<br>**(n=67)** |  | **F**<br>**(n=42)** |  | **M**<br>**(n=25)** |  |
| --- | --- | --- | --- | --- | --- | --- |
|  | **M** | **SD** | **M** | **SD** | **M** | **SD** |
| **Age (y)** | 37.2 | 13.5 | 40.5 | 13.5 | 31.7 | 11.7 |
| **BM (kg)** | 74.3 | 16.1 | 67.4 | 14.1 | 85.8 | 12.3 |
| **Height (cm)** | 170.2 | 9.3 | 165.2 | 7.1 | 178.5 | 6.1 |
| **BMI (kg/m<sup>2</sup>)** | 24.6 | 4.9 | 23.9 | 4.9 | 25.7 | 4.7 |
| **FFMI (kg/m<sup>2</sup>)\*** | 18.5 | 2.8 | 16.9 | 1.8 | 21.3 | 1.9 |
| **BF%\*** | 26.4 | 10.2 | 30.2 | 9.3 | 20.2 | 8.3 |
| **FM (kg)\*** | 20.0 | 10.1 | 21.3 | 10.3 | 17.9 | 9.6 |
| **FFM (kg)\*** | 54.3 | 12.8 | 46.1 | 6.7 | 67.9 | 8.1 |
| **USG** | 1.018 | 0.006 | 1.017 | 0.006 | 1.021 | 0.006 |

### Reliability

ICC values for all outcomes were ≥0.993, with absolute TEMs of 0.8% for BF% and 0.6 kg for FM and FFM. Full reliability results are displayed in Table 2.

**Table 2.** Test-Retest Reliability of Hume Pod for Body Composition Assessment. Data from n=65 participants. *BF%: body fat percentage; FM: fat mass; FFM: fat-free mass; SD: standard deviation; TEM: technical error of measurement; LSC: least significant change; ICC: intraclass correlation coefficient*.

|  | **BF%** | **FM (kg)** | **FFM (kg)** |
| --- | --- | --- | --- |
| **Mean (Test 1)** | 26.5 | 19.7 | 54.3 |
| **SD (Test 1)** | 9.7 | 8.8 | 13.6 |
| **Mean (Test 2)** | 26.6 | 19.8 | 54.2 |
| **SD (Test 2)** | 9.5 | 8.7 | 13.3 |
| **Mean Difference** | -0.1 | -0.1 | 0.1 |
| **SD of Mean Difference** | 1.1 | 0.9 | 0.9 |
| **TEM (absolute)** | 0.8 | 0.6 | 0.6 |
| **TEM (relative)** | -- | 3.2 | 1.2 |
| **LSC** | 2.2 | 1.8 | 1.8 |
| **ICC** | 0.993 | 0.995 | 0.998 |

### Validity (4C Comparison)

BF% values did not differ between the 4C model ([M ± SD] 26.4 ± 10.2%) and the Hume Pod (27.1 ± 9.8%) when compared via *t*-test (t(66) = 1.6; *p*=0.11). Additionally, statistical equivalence in BF% values was observed (t(66) = -3.7; *p*<0.01). When examining the linear relationship between values (Figure 1A), neither the slope (95% CI: 0.90, 1.06) nor intercept (95% CI: -2.39, 2.11) of the line of best fit significantly differed from the line of identity. A very strong Pearson’s correlation (*r*=0.95; *p*<0.001) was noted, along with a MAE of 2.6%. Bland-Altman analysis (Figure 1B) indicated no proportional bias (95% CI for slope: -0.11, 0.05).

**Figure 1.**
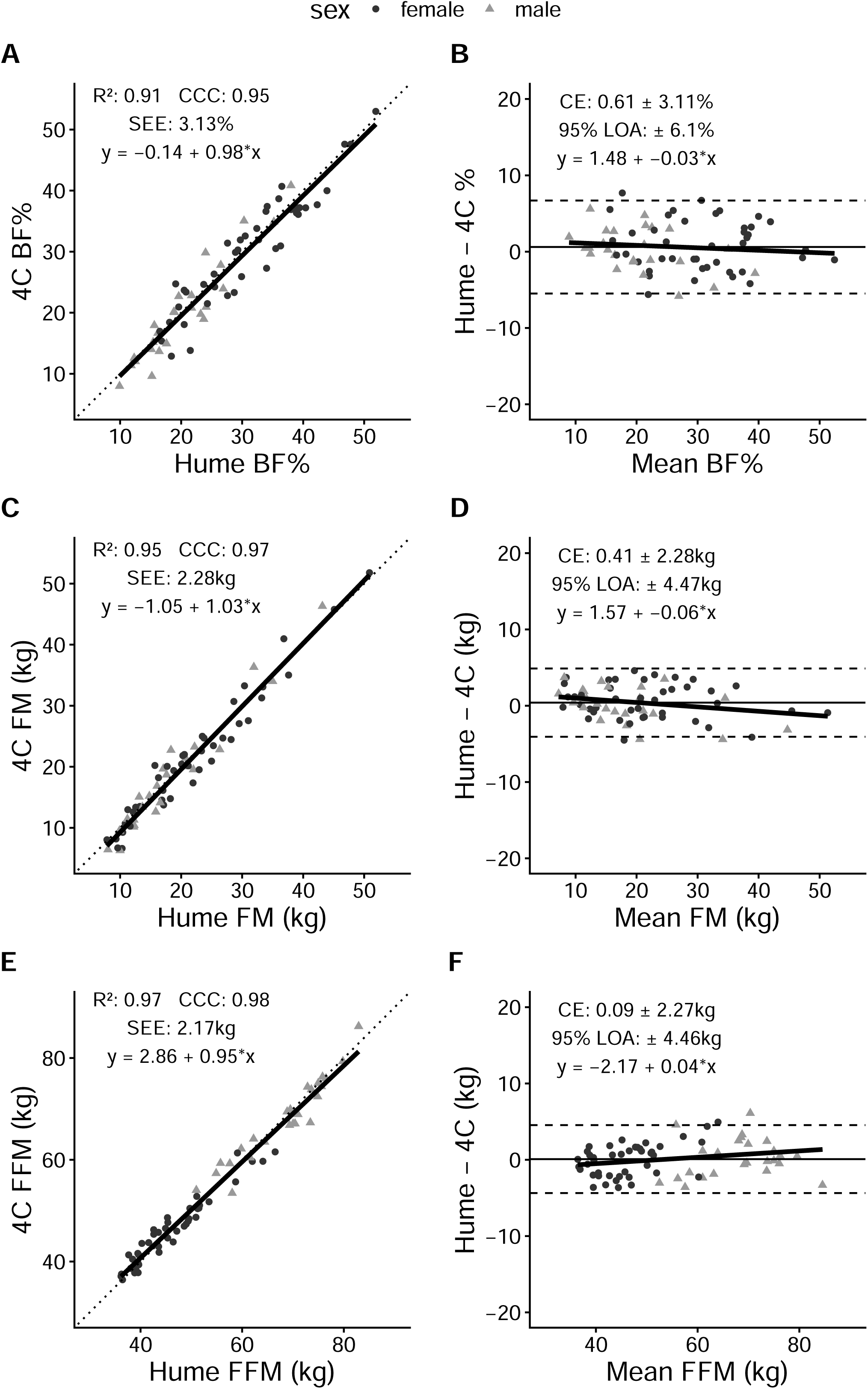
Validity of Hume Pod Compared to 4-compartment Model. Panels A, C, and E present line-of-identity plots for body fat percentage (BF%), fat mass (FM), and fat-free mass (FFM), respectively. The dotted line represents the line of identity (y = x), while the solid line represents the ordinary least squares regression line. The coefficient of determination (R^2^), standard error of the estimate (SEE), and linear equation for the line of best fit are displayed. Panels B, D, and F present Bland–Altman plots comparing Hume Pod estimates with the 4-compartment (4C) model. The solid horizontal line represents the constant error (CE), and the dashed horizontal lines represent the 95% limits of agreement (LOA). The slope of the displayed linear equation represents the degree of proportional bias present. Female participants are shown as circles and male participants as triangles.

FM values did not differ between the 4C model ([M ± SD] 20.0 ± 10.1 kg) and the Hume Pod (20.4 ± 9.5 kg) when compared via *t*-test (t(66) = 1.5; *p*=0.15). Additionally, statistical equivalence in FM values was observed (t(66) = -2.1; *p*=0.02). When examining the linear relationship between values (Figure 1C), neither the slope (95% CI: 0.97, 1.09) nor intercept (95% CI: -2.37, 1.09) of the line of best fit significantly differed from the line of identity. A very strong Pearson’s correlation (*r*=0.97; *p*<0.001) was noted, along with a MAE of 1.9 kg. Bland-Altman analysis (Figure 1D) indicated no significant proportional bias (95% CI for slope: -0.11, 0.00).

FFM values did not differ between the 4C model ([M ± SD] 54.3 ± 12.8 kg) and the Hume Pod (54.4 ± 13.4 kg) when compared via *t*-test (t(66) = 0.32; *p*=0.75). Additionally, statistical equivalence in FFM values was observed (t(66) = -9.4; *p*<0.01). When examining the linear relationship between values (Figure 1E), the slope of the best fit line was slightly lower than a value of 1 (95% CI: 0.91, 0.99), and the intercept was greater than 0 (95% CI: 0.62, 5.09). A very strong Pearson’s correlation (*r*=0.99; *p*<0.001) was noted, along with a MAE of 1.9 kg. Bland-Altman analysis (Figure 1F) indicated no proportional bias (95% CI for slope: 0.00, 0.08).

### Validity (DXA Comparison)

BF% values differed between DXA ([M ± SD] 28.9 ± 10.0%) and the Hume Pod (27.1 ± 9.8%) when compared via *t*-test (t(66) = -4.47; *p*<0.01). Additionally, statistical equivalence in BF% values was not observed (t(66) = 0.48; *p*=0.31). However, when examining the linear relationship between values (Figure 2A), the slope of the line of best fit did not differ from the line of identity (95% CI: 0.87, 1.04). In contrast, the intercept of the best fit line exceeded 0 (95% CI: 0.65, 5.39). A very strong Pearson’s correlation (*r*=0.94; *p*<0.001) was noted, along with a MAE of 3.0%. Bland-Altman analysis (Figure 2B) indicated no proportional bias (95% CI for slope: -0.10, 0.07).

**Figure 2.**
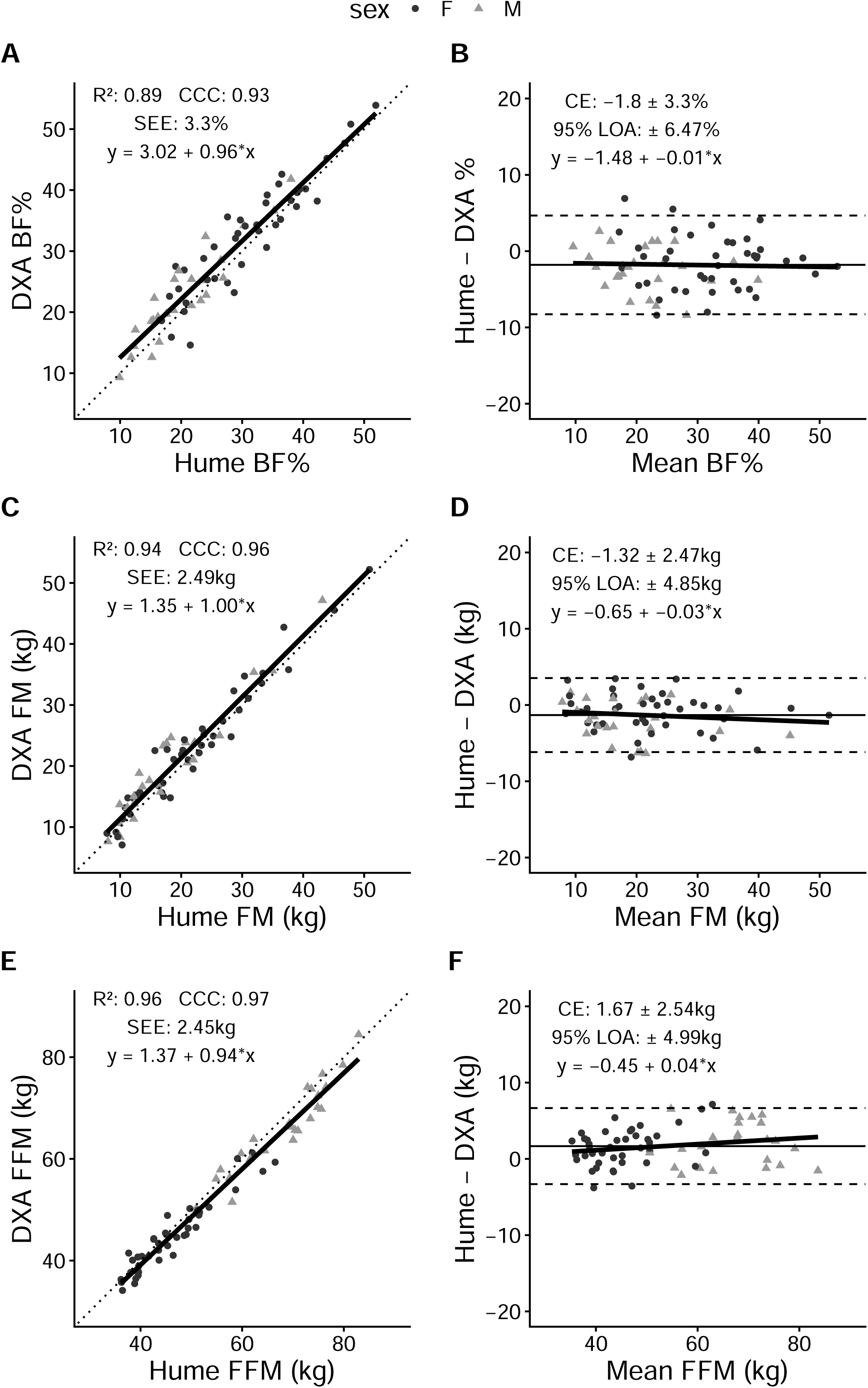
Validity of Hume Pod Compared to Dual-energy X-ray Absorptiometry. Panels A, C, and E present line-of-identity plots for body fat percentage (BF%), fat mass (FM), and fat-free mass (FFM), respectively. The dotted line represents the line of identity (y = x), while the solid line represents the ordinary least squares regression line. The coefficient of determination (R^2^), standard error of the estimate (SEE), and linear equation for the line of best fit are displayed. Panels B, D, and F present Bland–Altman plots comparing Hume Pod estimates with dual-energy X-ray absorptiometry (DXA). The solid horizontal line represents the constant error (CE), and the dashed horizontal lines represent the 95% limits of agreement (LOA). The slope of the displayed linear equation represents the degree of proportional bias present. Female participants are shown as circles and male participants as triangles.

FM values differed between DXA ([M ± SD] 21.7 ± 9.8 kg) and the Hume Pod (20.4 ± 9.5 kg) when compared via *t*-test (t(66) = -4.38; *p*<0.01). Additionally, statistical equivalence in FM values was not observed (t(66) = -0.79; *p*=0.78). However, when examining the linear relationship between values (Figure 2C), neither the slope (95% CI: 0.93, 1.06) nor intercept (95% CI: -0.09, 2.80) of the line of best fit differed from the line of identity. A very strong Pearson’s correlation (*r*=0.97; *p*<0.001) was noted, along with a MAE of 2.2 kg. Bland-Altman analysis (Figure 2D) indicated no proportional bias (95% CI for slope: -0.10, 0.03).

FFM values differed between DXA ([M ± SD] 52.7 ± 12.9 kg) and the Hume Pod (54.4 ± 13.4 kg) when compared via *t*-test (t(66) = 5.38; *p*<0.01). However, statistical equivalence in FFM values was also observed (t(66) = -3.1; *p*<0.01). When examining the linear relationship between values (Figure 2E), the slope of the best fit line was slightly lower than a value of 1 (95% CI: 0.90, 0.99), but the intercept did not differ from 0 (95% CI: -1.16, 3.89). A very strong Pearson’s correlation (*r*=0.98; *p*<0.001) was noted, along with a MAE of 2.5 kg. Bland-Altman analysis (Figure 2F) indicated no proportional bias (95% CI for slope: -0.01, 0.09).

## Discussion

The purpose of the present investigation was to evaluate the test-retest reliability and validity of the Hume Pod, a popular consumer-grade bioelectrical impedance analyzer, relative to both a criterion 4C model and DXA. Overall, the Hume Pod demonstrated excellent agreement with the 4C model for estimates of BF%, FM, and FFM. Group-level differences between methods were small, statistical equivalence was observed for all three outcomes, and strong linear relationships were evident with no evidence of systematic or proportional bias. Comparison with DXA likewise demonstrated strong correlations and agreement, although statistically significant mean differences were observed between methods, as well as statistical equivalence only for FFM. Collectively, these findings suggest that the Hume Pod ranks among the more accurate consumer-grade bioelectrical impedance analyzers evaluated to date^(11)^ and support its potential utility for body composition assessment in settings where laboratory methods are unavailable.

Perhaps the most notable finding was the degree of agreement observed between the Hume Pod and the criterion 4C model. BF% varied between methods by only 0.6% on average, with a MAE of 2.6%, R^2^ of 0.91, and a Lin’s CCC of 0.95. Similarly, FM and FFM demonstrated CCCs of 0.97 and 0.98, respectively, with mean absolute errors <2 kg. The regression slopes for BF% and FM were statistically indistinguishable from the line of identity, while Bland-Altman analyses demonstrated no evidence of proportional bias. Our lab’s prior work comparing fourteen consumer-grade bioimpedance analyzers to a similar 4C model presents a unique opportunity to contextualize the Hume Pod’s performance.^(11)^ In this investigation, the validity of BF% estimates ranged from poor to very good, despite the apparent similarity of the tested devices. For example, BF% SEE values ranged from 3.2% to 7.5%, with 95% LOAs ranging from ±6.6% to ±14.7%. The two top performing consumer analyzers for cross-sectional validity were the InBody H2ON and Tanita BC-568 InnerScan. Similar to the Hume Pod, both devices use contact electrodes for both hands and both feet. The R^2^, CCC, SEE, and 95% LOA when comparing each analyzer’s BF% estimates to the 4C model were as follows – InBody H2ON: R^2^ of 0.88, CCC of 0.94, SEE of 3.2%, and LOA of ±6.6%; Tanita BC-568: R^2^ of 0.81, CCC of 0.89, SEE of 4.0%, and LOA of ±8.2%. In the present investigation, the Hume Pod demonstrated a R^2^ of 0.91, CCC of 0.95, SEE of 3.1%, and LOA of ±6.1% for BF%. Therefore, the Hume Pod results were similar but slightly superior to the InBody H2ON, which previously ranked as the consumer device with the highest validity for cross-sectional assessments. Most of the equipment and laboratory procedures were identical between our two studies, improving the ability to directly compare results across studies. Collectively, our results indicate that the cross-sectional validity of the Hume Pod is superior to numerous previously tested consumer bioimpedance analyzers.

The present findings reinforce an important observation from our previous consumer bioimpedance research, namely that performance varies considerably across apparently similar individual devices. Many of the tested analyzers exhibited very similar design and hardware characteristics, suggesting that consumers would not readily be able to distinguish reasonably accurate devices from those that perform poorly. These observations emphasize that the validity of one specific consumer bioimpedance analyzer should not be generalized to other commercially available products, even when similar electrode configurations or measurement principles are employed. Rather, device-specific validation remains essential because prediction algorithms, signal processing methods, and proprietary analytical approaches differ substantially among manufacturers.^(7; 10)^ This consideration is increasingly important as consumer body composition analyzers with progressively more sophisticated analytical capabilities are developed and incorporated into health and fitness decision-making.

Although agreement between the Hume Pod and DXA was also strong, small systematic differences between devices were observed for BF%, FM, and FFM. Interestingly, the Hume Pod is advertised as being validated against DXA.^(13)^ Based on accuracy reports on the manufacturer’s website,^(13)^ which were seemingly provided by a contracted laboratory, the mean FM values in a sample of 50 individuals were 17.8 kg for DXA and 16.0 kg for the Hume Pod (∼1.8 kg mean underestimation by the Hume Pod). Although the model of DXA scanner is not listed, this magnitude is notably similar to the mean underestimation of 1.3 kg observed in the present study. The reported correlation between the Hume Pod and DXA for FM reported by the manufacturer was 0.97, corresponding to a R^2^ of 0.94, assuming the correlation being reported was Pearson’s *r*. This R^2^ value precisely matches the R^2^ in the present investigation. In contrast, the 95% LOA were reported by the manufacturer as 4.185, -0.454, corresponding to a total LOA width of ∼4.64 kg. In the present study, the observed LOA between DXA and Hume Pod FM were ±4.85 kg, corresponding to a total LOA width of ∼9.7 kg. This notable difference could potentially be attributable to truly lower variation in errors in the sample of individuals investigated by the manufacturer, although it is also possible that a difference in calculation method or accidental mis-presentation of the metric occurred. Overall, excluding the LOA values, the FM comparisons between DXA and Hume Pod are notably similar between the manufacturer’s report and the present independent investigation. Importantly, the transparency of the manufacturer to present their validation results publicly on their website allowed for the present comparisons. The consumer-grade body composition assessment industry would benefit substantially if more manufacturers similarly pursued validation and transparently shared their results in this manner.

The slightly varying performance of the Hume Pod when compared to the 4C model vs. DXA likely reflects, at least in part, known methodological differences between DXA and multi-compartment body composition models. Relative to the 4C model, DXA relies on more assumptions regarding the composition and hydration of the FFM that may not hold equally across all individuals.^(15; 24)^ Consequently, systematic differences between DXA and multi-compartment models have been documented previously, particularly among individuals differing in age, sex, adiposity, or hydration status.^(25; 26)^ Given that the Hume Pod generally demonstrated excellent agreement with the 4C model while showing strong performance but slight differences from DXA, the present findings support the importance of interpreting validation studies within the context of the chosen reference method rather than assuming all laboratory methods represent identical targets.^(27)^

The reliability analyses in the present investigation support the potential utility of the Hume Pod for repeated assessments; however, importantly, the reliability analysis was based on short-term test-retest measurements and therefore reflects technical error rather than biological error.^(28; 29)^ ICCs ranged from 0.993 to 0.998 across outcomes, indicating excellent short-term repeatability. The observed TEMs were small (0.8% for BF% and 0.6 kg for FM and FFM). While these TEM values indicate that a change in FM or FFM of >0.6 kg would be needed to exceed the observed technical error, it is expected that between-day reliability assessments – which include the added component of biological error – would result in larger errors. Indeed, a recent investigation from our laboratory indicated that bioimpedance analyzers employing contact electrodes demonstrated greater biological error than technical error when within- and between-day reliability assessments were performed.^(28)^ As compared to our prior investigation of consumer bioimpedance devices, which similarly included test-retest reliability assessments within the same session, the present TEM of 0.8% for the Hume Pod BF% was slightly higher than the TEMs for previously evaluated consumer devices, which were all <0.5%.^(11)^ Interestingly, this indicates that slightly higher technical error may be present for the Hume Pod as compared to previously evaluated analyzers, despite its excellent validity performance. In both studies, TEM values primarily reflect the technical precision of the analyzer in laboratory conditions, the positioning of the participant, and any unknown software features that may be present when repeated assessments are performed. Accordingly, within-day reliability results should not be interpreted as the minimum detectable physiological change during routine home use, where additional biological variation arising from hydration status, dietary habits, physical activity, and normal day-to-day variability would be expected to increase measurement error.^(29; 30; 31)^ Nevertheless, the generally high repeatability observed under controlled conditions indicates that the analyzer itself contributes relatively little technical variability to repeated measurements, consistent with our previous work on consumer bioimpedance.^(11)^

The present investigation had several strengths. First, validation was performed relative to both a 4C model and DXA. The 4C model comparison provides arguably the most rigorous evaluation of the Hume Pod, while the DXA represents a common standard in the development of consumer body composition tools and allowed direct comparison to manufacturer-promoted validation results. Second, comprehensive statistical evaluation included multiple complementary assessments of validity and reliability, allowing for a thorough understanding of the Hume Pod’s performance. Finally, the inclusion of participants spanning a broad range of body size and composition improves the generalizability of the findings within generally healthy adults.

Several limitations of the present work should also be considered. Participants were generally healthy adults, and the present findings may not extend to pediatric populations, older adults, individuals with severe obesity, or patients with medical conditions known to alter fluid distribution. Additionally, as discussed, reliability was assessed using repeated measurements obtained during a single laboratory visit and therefore primarily reflects technical precision rather than day-to-day reproducibility. Future investigations should evaluate between-day reliability under laboratory and typical home-use conditions and examine the longitudinal validity of the Hume Pod for tracking changes in body composition during weight loss, exercise training, and clinical interventions. Such work will hold great practical importance since many consumers use these devices primarily to monitor changes over time rather than to obtain a single estimate of body composition.

Overall, the present independent study indicates that the Hume Pod demonstrates good reliability and strong agreement with a criterion 4C model for estimating body composition in healthy adults. Its validity compares favorably to numerous other consumer-grade bioelectrical impedance analyzers evaluated to date. While additional work is warranted to establish longitudinal validity and performance across broader clinical populations, the present findings support the Hume Pod as an option for consumer body composition assessment when laboratory methods are unavailable.

## Data Availability

Data produced in the present study may be available upon reasonable request to the authors.

## List of abbreviations

4C: 4-compartment
BF%: body fat percentage
BIA: bioelectrical impedance analysis
BM: body mass
BV: body volume
CCC: concordance correlation coefficient
CI: confidence interval
DXA: dual-energy X-ray absorptiometry
FFM: fat-free mass
FM: fat mass
ICC: intraclass correlation coefficient
LOA: limits of agreement
MAE: mean absolute error
Mo: bone mineral
SEE: standard error of the estimate
TBW: total body water
TEM: technical error of measurement.

## Acknowledgements

The authors wish to acknowledge the participants for their efforts as part of this study.

## Authorship

Conceptualization: GMT, CMV, CMF; Data curation: CMV, CMF, AEW, MHS, JAW, RAR, JRA, AM; Formal Analysis: GMT; Investigation: GMT, CMV, CMF, AEW, MHS, JAW, RAR, JRA, AM; Methodology: GMT, CMV, CMF; Project administration: GMT, CMV, CMF, AEW, MHS; Resources: GMT; Supervision: GMT; Writing – original draft: GMT; Writing – review & editing: GMT, CMV, CMF, AEW, MHS, JAW, RAR, JRA, AM

